# Development and validation of diagnostic and prognostic prediction tools for dental caries in young children: A protocol

**DOI:** 10.1101/2025.05.14.25327586

**Authors:** Yeganeh Khazaei, Saritha Kodikara, Catherine Butler, Nicole L Messina, Kim-Anh Lê Cao, Stuart Dashper, Mihiri Silva

**Affiliations:** Melbourne Dental School, University of Melbourne, Melbourne, Victoria, Australia; Melbourne Integrative Genomics, School of Mathematics and Statistics, University of Melbourne, Melbourne, Victoria, Australia; Inflammatory Origins, Murdoch Children’s Research Institute, Melbourne, Victoria, Australia; Department of Paediatrics, The University of Melbourne, Parkville, Victoria, Australia; Infectious Diseases Group, Murdoch Children’s Research Institute, Parkville, Victoria, Australia

**Author notes:** Corresponding author: Mihiri Silva, 720 Swanston St, Carlton VIC 3053, Australia.

## Abstract

**Introduction:** Dental caries is the most common oral disease worldwide, affecting up to 90% of children globally. It can lead to pain, infection, and impaired quality of life. Early prevention is a key strategy for reducing the prevalence of dental caries in young children. Valid and reliable diagnostic or prognostic tools that enable accurate individualised prediction of current or future dental caries are essential for facilitating personalised caries prevention and early intervention. However, no efficacious tools currently exist in early childhood—the optimal period for disease prevention. We aim to develop and validate diagnostic and prognostic prediction tools for dental caries in young children, utilising a combination of environmental, physical, behavioural and biological early life data.

**Methods and analysis:** Data sources include two prospective studies, with a total sample size of approximately 600 children. These cohorts have collected detailed demographic, antenatal, perinatal and postnatal data from medical records and parent-completed questionnaires and biological samples including a dental plaque swab. Candidate predictor variables will include sociodemographic characteristics, health history, behavioural, and microbiological characteristics. The outcome variable will be the presence, incidence, or severity of dental caries diagnosed using the International Caries Detection and Assessment System (ICDAS). Statistical and machine learning approaches will be utilised for selection of predictor variables and model development. Internal validation will be conducted using resampling methods (i.e. bootstrapping) and nested cross validation. Model performance will be evaluated using standard performance metrics such as accuracy, discrimination, and calibration. Where feasible, external validation will be performed in an independent cohort. Model development and reporting will be guided by the Transparent Reporting of a multivariable prediction model for Individual Prognosis or Diagnosis (TRIPOD) statement and the Prediction model Risk Of Bias Assessment Tool (PROBAST) guidelines.

**Discussion:** This protocol describes data collection procedures, outcome and predictor variable configuration, and planned omics-based diagnostic and prognostic prediction analyses. The study employs a discovery-driven approach for the development and validation phases, allowing findings from select steps to inform subsequent stages.

**Ethics and dissemination:** This study has ethical and governance approval from The Royal Children’s Hospital Melbourne Human Research Ethics Committee (HREC/111803/RCHM-2024).

**Strengths and limitations of this study:**

- Analysis of two rare, prospectively collected datasets from early childhood, comprising longitudinal and cross-sectional early-life questionnaire data, oral microbiome profiles, and clinical dental assessments.
- Use of a gold-standard dental caries measurement based on ICDAS, providing a validated, standardised, and detailed evaluation of caries status.
- Rigorous study design supported by a comprehensive and pre-specified data analysis plan.
- Variability in early-life questionnaire data collection across cohorts may introduce heterogeneity, potentially affecting the comparability and transportability of the prediction models.
- The lack of external validation in an independent dataset beyond the research setting may limit the generalisability of the findings to broader paediatric populations.

## INTRODUCTION

Dental caries is among the most common health issues worldwide, affecting 514 million children.^1^ In Australia, 42% of children aged 5–10 have experienced dental caries in their primary teeth.^2^ The economic burden of oral diseases exceeded AUD 11.1 billion in Australia in 2021–22 and reached ∼USD 710 billion globally in 2019.^2,3^ Early Childhood Caries (ECC), treated or untreated dental caries in a child under the age of six years is a global health problem.^4,5^ ECC leads to substantial health, social and economic burdens on children, families, healthcare infrastructure, and the community at large.^6^ ECC can lead to pain, infections, and difficulties in eating, speaking, and sleeping, ultimately hindering school attendance, academic performance, and self-esteem.^7,8^

Dental caries is a chronic disease arising from complex interactions over time between the oral microbiota, host susceptibility at the tooth, mouth, and individual levels, and environmental factors.^9^ A key cause of caries is frequent sugar consumption—which triggers an imbalance (dysbiosis) in the normal oral microbiota, the central mechanism in caries pathogenesis. This dysbiosis manifests as an overgrowth of certain cariogenic microorganisms, causing demineralization or destruction of susceptible tooth surfaces.^10,11^ Dysbiosis in early childhood—particularly the first 2000 days of life—may also affect lifelong oral microbiota trajectory.^12^ This period is characterized by significant developmental plasticity, during which a child‘s oral and overall health is highly responsive to environmental, nutritional, and social exposures.^13^ Studies suggest that certain exposures in this window—such as birth mode, feeding practices, and oral hygiene—can modify the acquisition and development of the oral microbiota, thereby shaping caries risk.^12,14^ Additionally, distinct differences in oral microbial composition have been observed between caries-free and caries-active children, with microbial alterations detectable up to 24 months before clinical manifestations of caries.^15^ Hence, dysbiosis during this period is a critical determinant of subsequent dental caries and may predict children likely to develop severe ECC.^16^

Early detection of ECC is a key strategy to allow preventive interventions in young children.^16^ Valid and reliable tools that accurately predict the presence or severity of current (diagnostic) or future (prognostic) caries can support triage in primary care settings and help personalise preventive interventions. This includes both individual behavioural change support and therapeutic procedures to prevent the development of clinical disease. Such tools can support investment of limited resources for individuals at greatest need.^17^ Early-life factors that have been identified as predictive of ECC can be utilised in diagnostic and prognostic prediction tools for young children.^9,18^ Additionally, as oral microbial dysbiosis precedes clinical disease, incorporating microbiome data, combined with early-life factors, may improve their accuracy.^19,20^

Although some tools exist for specific settings and populations, reliable and validated tools for Australian children are lacking.^21^ Most existing tools mainly function as caries risk assessment tools to evaluate and identify current caries risk factors, rather than as predictive models that enable healthcare professionals to accurately predict ECC outcomes.^22,23^ The most frequently validated tools provide only moderate to weak evidence regarding their reliability, validation, and standardization, with inappropriate statistical methods and limited data to support their use in paediatric populations.^24,25^ Moreover, these tools have relied primarily on established clinical disease indicators to predict future risk—manifestly too late for meaningful primary prevention.^18^ Additionally, despite their potential, microbiome-based tools are impeded by small sample sizes, lack of standardized outcome measures, limited taxonomic resolution, and poor reporting. Niche study populations and minimal emphasis on external or temporal validation further restrict broader clinical implementation.^20^

In line with the recommendations to publish protocols according to the Transparent Reporting of a multivariable prediction model for Individual Prognosis or Diagnosis (TRIPOD) statement and the Prediction model Risk Of Bias Assessment Tool (PROBAST) guidelines^26-28^, this discovery-driven study protocol pre-specifies the predictor variables and analytical plan for the development and validation of diagnostic and prognostic prediction tools for dental caries in young children.

### Research hypotheses

Early life exposures can accurately diagnose and predict ECC. By identifying microbial biomarkers that can reflect current or future caries status, the microbiome can be used as a component of an ECC prediction model to improve accuracy.

### Study aims and objectives

The overall aim of the study is to develop and validate tools to diagnose and predict ECC, utilising a combination of early-life data with oral microbiome data, to be used in primary care settings.

This study includes the following aims:

I. To develop and internally validate ***diagnostic prediction*** models for ECC using; (a) questionnaire-based data (b) microbiome data (c) a combination of both.
II. To develop and internally validate ***prognostic prediction*** models for ECC using; (a) questionnaire-based data (b) microbiome data (c) a combination of both.
III. To evaluate the change in predictive power of diagnostic and prognostic prediction models for ECC in young children with and without a microbiome component.

## METHODS AND ANALYSIS

### Data sources

This study will utilise cross-sectional and longitudinal data from two prospectively collected studies, MIS BAIR and Infant2Child.

#### “Melbourne Infant Study – BCG for Allergy and Infection Reduction (MIS BAIR)” (HREC 33025)

MIS BAIR was initially established in 2013 as a phase III, multicentre RCT which included 1272 neonates (birth to ten days of life) followed-up to 5 years (5y) of age.^29^ The RCT aimed to evaluate the effect of neonatal BCG (tuberculosis) vaccination on allergy (including eczema and atopic sensitisation), infection, and asthma over the first 5y of life. Birth-hospital medical records, biological samples, parent-completed online questionnaires and study visits (1y and 5y) have been used to collect detailed longitudinal social and health data throughout the study. Parents completed online questionnaires 3 monthly in the first year and 6 to 12 monthly from 1 to 5y of age (Fig. 1). At 5y of age, an additional, optional, dental cohort study was embedded within the RCT, and participants were invited to complete additional surveys and participate in a dental examination for measurement of dental caries and collection of a dental plaque sample (n=219).

**Figure 1.**
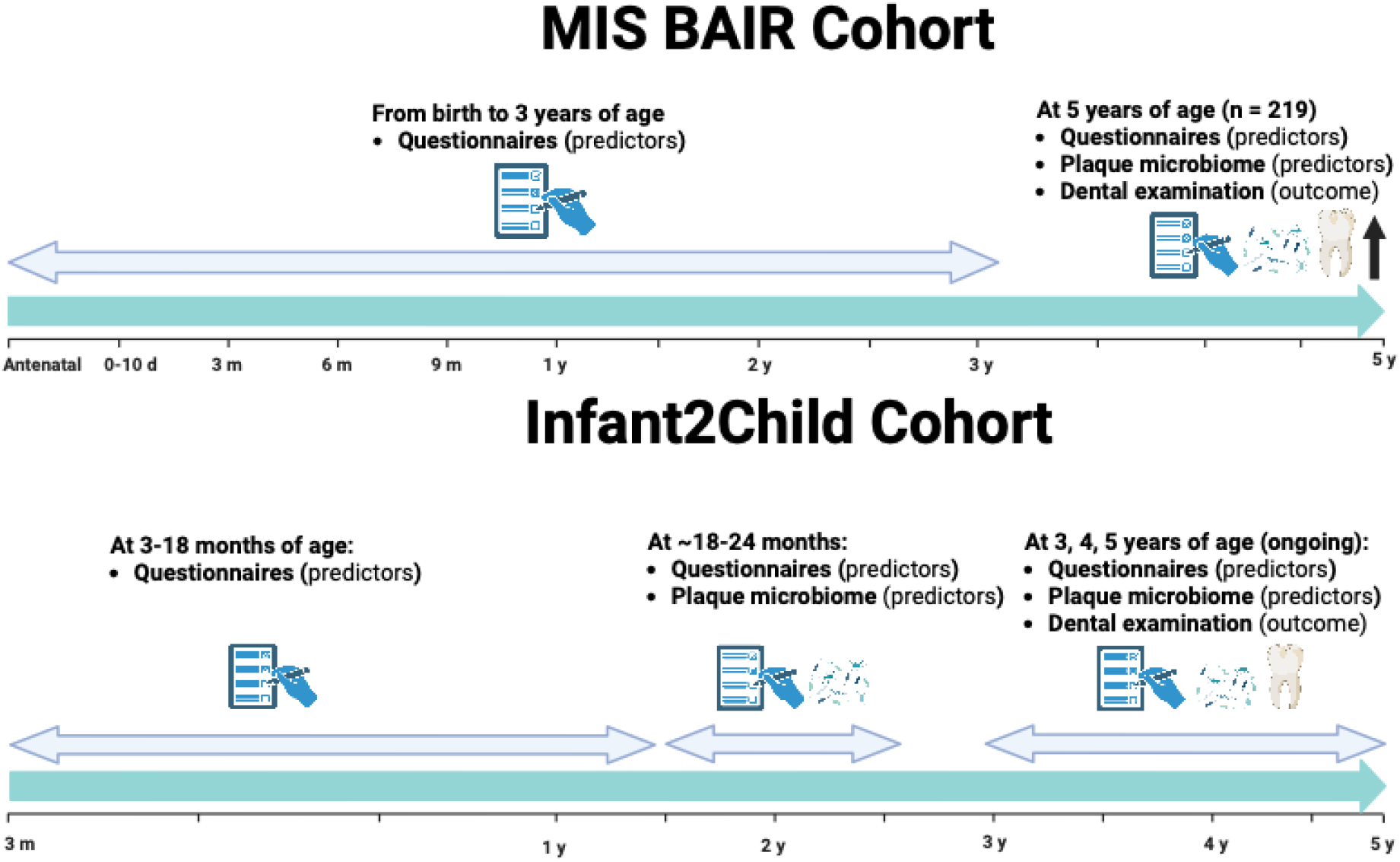
Cohorts and data collection schedule (Created in BioRender. Khazaei, Y. (2025) https://BioRender.com/gy2b19r)

#### “Infant2Child (I2C): Optimising nutrition in early life to reduce childhood dental caries” (HREC 80506)

Established in 2021, I2C is an ongoing longitudinal cohort. Data from 3-18 months of age have been obtained through parent-completed online questionnaires. At 2y of age, in addition to parent-completed online questionnaires, an at-home dental plaque swab is collected. At 3, 4, and 5y of age data is collected through questionnaires, and dental assessments (including measurement of dental caries and dental plaque sample collection) (Fig. 1). Data collection is ongoing, with an anticipated total sample size of approximately 400 children.

### Inclusion criteria

In MIS BAIR, we will analyse data from participants who underwent dental examinations at 5 years of age. In I2C, we will include participants who underwent dental examinations at a minimum of one timepoint (3, 4, or 5y of age).

### Selection of potential questionnaire-based predictors

To identify potential candidate predictors for caries in young children, we conducted a targeted scoping review ^30^ and consulted knowledge users in public health, along with expertise from our team. This systematic approach was utilised to refine several available variables into a concise set of core predictors—covering sociodemographic and health history, behavioural, and microbiological risk factors. The availability of core predictor variables in the MIS BAIR and Infant2Child data sets is outlined in Table 1 (Supplementary Material A, Fig. A1).

**Table 1.** Predictor variables.

| Category | Subcategory | MIS BAIR | I2C |
| --- | --- | --- | --- |
| Sociodemographic/Health History | Family and demographic | ✓ | ✓ |
|  | Medical health | ✓ | ✓ |
|  | Household environment | ✓ |  |
|  | Maternal prenatal data | ✓ |  |
|  | Birth data | ✓ |  |
|  | Oral health-related quality of life* |  | ✓ |
| Behavioural | Infant feeding | ✓ | ✓ |
|  | Dental habits | ✓ | ✓ |
|  | Diet | ✓ | ✓** |
|  | Fluoride exposure | ✓ | ✓ |
| Microbiological | Plaque samples | ✓ | ✓ |

\* Parental-Caregiver Perceptions Questionnaire (P-CPQ)<sup>31</sup>
\*\* using INFANT study food frequency questionnaires (FFQ)<sup>32</sup>

### Plaque microbiome laboratory procedures and bioinformatics

All laboratory analytical procedures will be completed without knowledge of caries status. Samples will be identified only by date, examination number, and subject ID. Mechanical and chemical genomic DNA (gDNA) extraction, and purification of samples will be performed.^16^ DNA sequencing of the full-length 16S rRNA gene will be conducted using high-fidelity long-read sequencing by Pacific Biosciences (PacBio) Sequel technology.^33^ A detailed description of bioinformatics is provided in Supplementary Material A.

### Outcome

#### Clinical examination

Dental examinations are conducted by trained and calibrated registered dental practitioners. Dental caries experience is recorded for each tooth surface using the two-digit International Caries Detection and Assessment System (ICDAS) criteria (Supplementary Material B, Table B1).^34^ ICDAS data are recorded on a custom-built dental chart within the Research Electronic Data Capture (REDCap; Vanderbilt University) system hosted at MCRI.^35^

#### Outcome variable configuration

Our proposed outcomes include binary, categorical, count, and time-to-event. To configure ICDAS scores to a dmft (decayed, missing, filled surfaces) score, we will apply severity-specific thresholds, as outlined in Table 2.

**Table 2.**
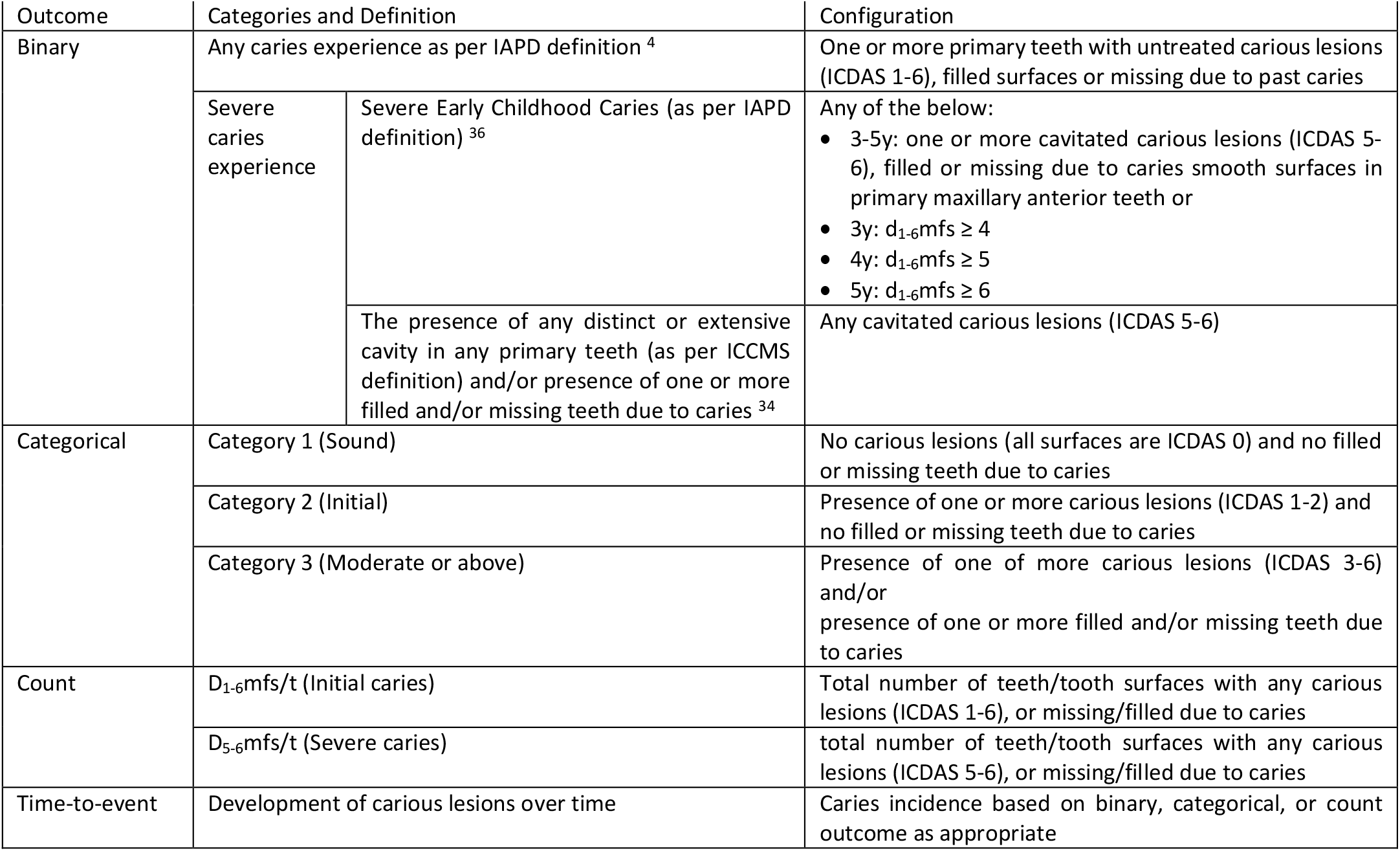
Outcome variable configuration.

### Sample size considerations

We conducted a power analysis in response to recent calls for rigorous justification in prediction modelling research.^37^ As the sample size for our initial analysis in the existing MIS BAIR cohort is fixed, we performed a post-hoc power analysis to determine the maximum number of parameters that can be included in the model, to avoid overfitting.^38^ Using the pmsampsize package in R,^39^ we followed the criteria outlined by Riley et al.,^40,41^ which accounts for an adequate number of events per predictor parameter, outcome proportion in the study population, the expected predictive performance of the model, and model complexity and overfitting.

We performed an initial exploratory analysis of MIS BAIR data and determined an outcome prevalence of ∼14% for Severe-ECC. We assumed a binary outcome with an expected predictive performance Area Under the Curve (AUC) of ∼80% —based on comparable risk prediction tools^42-47^—and a shrinkage factor of 90%. Under these parameters, we can include approximately 4-5 predictors in the model, anticipating about 33 events (with a 14% outcome prevalence) and yielding an events-per-predictor-parameter ratio of 8.09. For each potential outcome variable, we will select an appropriate regression model including multivariable binary logistic regression for binary outcomes, ordinal logistic regression for ordinal outcomes, or negative binomial regression for skewed count outcomes.^48^

This is a conservative estimate as it reflects a single cross-sectional timepoint based on early life questionnaire-based data. It does not account for the increased power afforded by longitudinal data, which would be obtained from multiple timepoints from both the MIS BAIR and I2C cohorts. The expected sample size of our ongoing I2C study (n=∼400) surpasses the calculated minimum estimates above. As a substantial body of evidence discourages data splitting,^38,49-51^ we will use both cohorts’ data for model development, with resampling methods for internal validation.^50^

In total, 219 oral plaque microbiome samples are expected from the MIS BAIR cohort. Additionally, the I2C cohort is expected to provide ∼700 samples across the four early-life timepoints at 2, 3, 4, and 5y. Based on the above calculations and previous studies, this sample size is predicted to be sufficient for our analyses.^42,43,45,47,52^

### Statistical analysis

All analyses will be performed using R Statistical Software.^53^ Model development will involve a series of predefined steps: (1) data exploration and pre-processing, (2) model building and internal validation, (3) and external validation. Figure 2 illustrates the complete workflow from model development steps through included datasets in each model that will be applied consistently across all models, separately for MIS BAIR and I2C. Note that the process is explorative, dynamic, and iterative between steps; outcomes of select steps will inform the final direction of model building.

**Figure 2.**
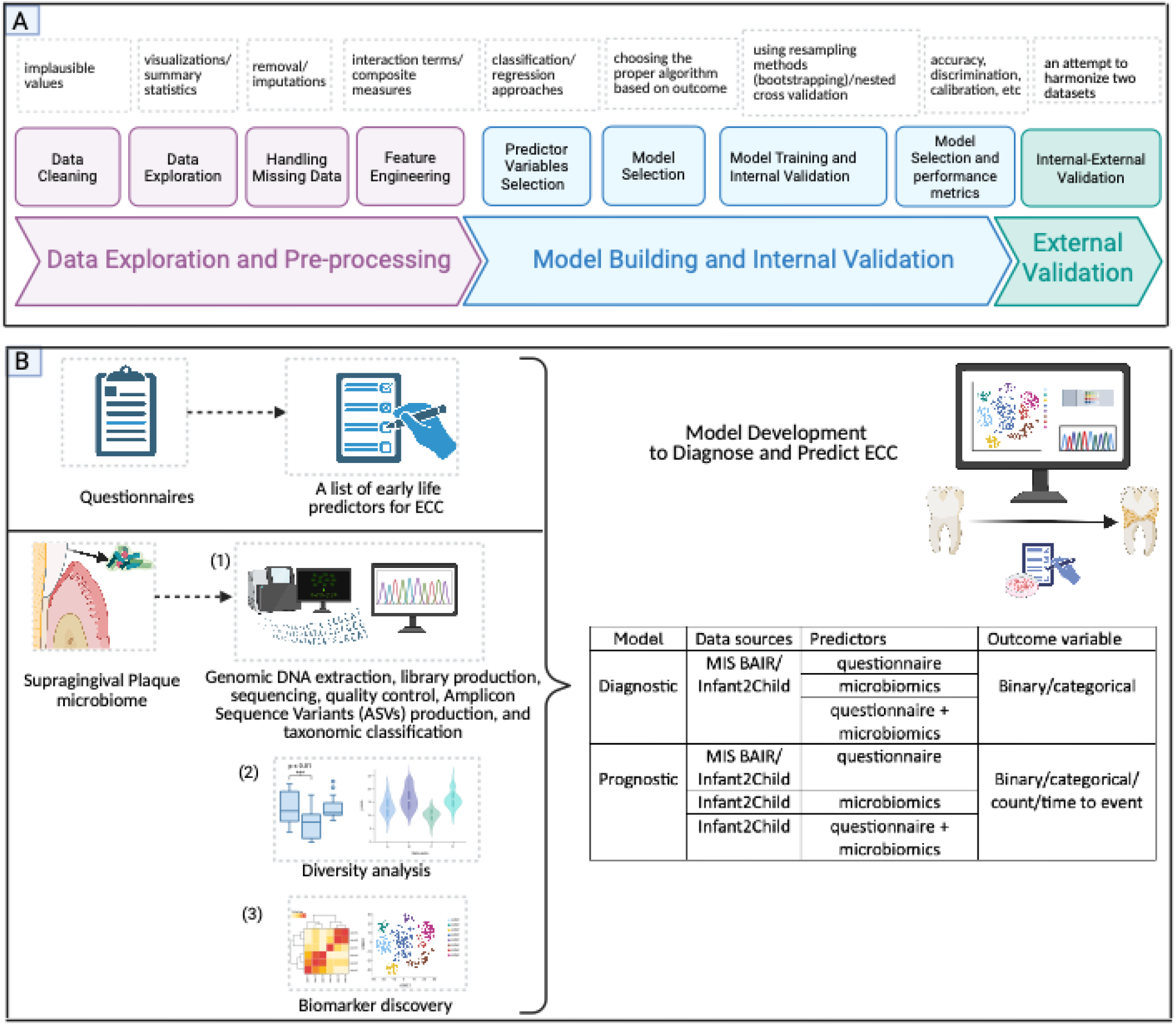
Pipeline diagram for (A) model development workflow, and (B) questionnaire and plaque microbiome as sources for predictor variables and a list of prediction models intended to be developed. (Created in BioRender. Khazaei, Y. (2025) https://BioRender.com/pkqlj57)

#### (1) Data exploration and pre-processing

##### Early life questionnaire data

Data cleaning for the MIS BAIR and I2C cohorts will be conducted before examining relationships between predictors and outcome. Specifically, the datasets will be reviewed for inconsistencies, errors, and anomalies (e.g., duplicate entries, implausible values) and corrected as needed. Descriptive statistics including mean, median, standard deviation, and interquartile range will be used to summarise continuous variables, and visualisations such as histograms and boxplots will be used to illustrate their distributions. Frequencies and proportions of categorical variables will be displayed using bar plots.^54^

We will investigate missing data in all potential predictors by examining both the pattern and mechanism of missingness. We may consider excluding predictors with a high proportion of missing values (e.g., more than 50% missingness), particularly when there is limited evidence of their predictive utility. Most importantly, we will always consider conducting multiple imputations by chained equations (MICE),^55,56^ incorporating the full set of predictors, the outcome variable, and relevant auxiliary variables (i.e., variables that are not direct predictors but can inform the imputation process).^57^

A list of predictors generated from these steps will then be used in variable selection and model development steps. We have provided more details and examples including feature engineering in Supplementary Material C1-C2.

##### Microbiome data

We will conduct a comprehensive plaque microbiome analysis in both cohorts to detect the most abundant taxa and understand differences in bacterial diversity and community structure between children in different groups (binary or categorical) at each timepoint. Diversity, within sample (alpha diversity) and between samples (beta diversity), will be analysed between these groups. We will also calculate relative abundances by normalizing read counts to one. We will conduct Principal Coordinate Analysis (PCoA) and graphically represent the resulting beta diversity patterns. We will run Permutational Multivariate Analysis of Variance (PERMANOVA) on beta diversity measures, overall and pairwise comparisons, to determine statistical significance when comparing community composition between groups and also for microbial divergence among populations.^58^ We will perform the Benjamini-Hochberg False Discovery Rate correction to adjust p-values for multiple comparisons. We will then visualise the Principal Component Analysis (PCA) results for the entire cohort as well as for the caries groups separately. This approach will help manage the high-dimensionality of the longitudinal microbiome data.^59^

Next, we will investigate any differences in the abundances of individual taxa between our groups (binary or categorical), by utilising Differential Abundance Analysis (DAA) methods.^60^ This is to explore cariogenic oral bacterial taxa at each timepoint that may diagnose caries. This can be performed using simple univariate models or multivariate methods such as sparse partial least squares discriminant analysis (sPLS-DA),^59,61^ and the analysis of the composition of microbiomes with bias correction.^16,60,62^

#### (2) Model building and internal validation

In the following sections, we outline our approach to developing separate diagnostic and prognostic prediction models, as illustrated (Fig. 2).

##### Predictor variables selection

To identify the most influential predictors of caries in young children, we will employ multiple complementary techniques.^48,52^ Classification approaches, such as discriminant analysis, will help determine which variables best classify individuals or taxa into categories (binary or categorical outcomes).^63^ Regression approaches, such as negative binomial regression and elastic net regression, including its variations such as adaptive elastic net, combined with Cox proportional hazards models will be utilised for count and time-to-event outcomes to select relevant predictors from the dataset while accounting for multicollinearity and overfitting (Supplementary Material C3).^64^ Notably, some methods, such as random forests and nested cross-validation, can integrate variable selection during model development.^65^

##### Diagnostic prediction models for caries in young children utilising (sections D1-3)

###### D1-Questionnaire-based data

We will explore the feasibility of developing a diagnostic prediction model using cross-sectional MIS BAIR and I2C early life data to concurrently diagnose caries in young children. We will train our model using resampling methods (i.e. bootstrapping) and nested cross validation,^52^ and we will compare multiple traditional statistical methods suitable for each outcome type (logistic regression, ordinal logistic regression) and machine learning algorithms (random forests or gradient boosting classifiers) to identify the best-performing approach for building our prediction model (in Supplementary Material C4, we have described the nested CV and the differences between these methods).^65^ Model selection will be based on:

1. performance metrics, including accuracy, sensitivity, specificity, F1-score, discrimination assessed by ROC-AUC, calibration, and overall performance by Brier score.
2. model complexity and clinical interpretability criteria by balancing between simpler, interpretable models such as logistic regression, and more complex, high-performing models including random forests and gradient boosting.
3. robustness and generalisability^65^

###### D2-Oral microbiome data

Utilising the results from data exploration and pre-processing of microbiome data, we will develop diagnostic prediction models. This will involve training a cross validated model (e.g., random forest, gradient boosting classifier) with the relative abundance of the overall microbiota composition, or the top-ranking important taxa, to cross sectionally diagnose caries. The choice of appropriate model selection based on the outcome type and resampling approach will follow section D1 (Supplementary Material C5).

###### D3-Combination of questionnaire and oral microbiome data

Ultimately, we will examine whether combining early-life predictors from section D1 and top-ranking bacterial taxa from section D2 improves diagnostic prediction of caries in young children. Specifically, we will assess if bacterial taxa can be effectively grouped into clusters and determine how the inclusion of early-life predictors affects model accuracy. Statistical analyses will follow previous methods. We will evaluate model performance and quantify any added predictive benefit from including microbiome data.

##### Prognostic prediction models for caries in young children utilising (sections P1-3)

###### P1-Questionnaire-based data

In sections P1-3, we outline the specific analytical methods employed to address repeated measurements over time and inherent correlations within subjects. These methods encompass both single-timepoint—when temporal variations in predictors and outcomes are not of primary interest—and longitudinal analyses involving data from multiple timepoints. Single-timepoint analyses will follow the same structured approach as per model development of diagnostic prediction models.

For longitudinal analyses, we will use mixed-effects models to accommodate both time-varying predictors, such as changes in dietary sugar intake reported through repeated questionnaires, as well as longitudinally collected outcomes. Different mixed-effects regression approaches will be selected according to outcome type (binary, categorical, count), using appropriate distributional assumptions and link functions. For instance, for repeated binary outcomes—such as the presence or absence of caries experience at each timepoint—we will use generalized linear mixed models (GLMMs) with a logit link functions to account for within-subject correlation.^66^ Time-to-event outcomes, such as the incidence of caries, will be explored using approaches including two-stage methods that incorporate estimated longitudinal parameters as covariates in Cox proportional hazards models. Joint models that simultaneously fit longitudinal and survival data will be an alternative as well.^67^ In parallel, we will explore machine learning approaches, including extensions of random forests and recurrent neural networks. These methods are effective in capturing complex, non-linear effects of time-varying exposures on caries and in detecting critical periods where certain exposure changes most influence caries development. All these methods will be compared in terms of their underlying assumptions, interpretability, model complexity, and suitability for the structure of our data.^68,69^

###### P2-Oral microbiome data

We will analyse the relative bacterial abundances at each timepoint to identify recurring dominant taxa.^16^ To capture temporal shifts in the oral plaque community, we will explore assigning bacterial taxa to clusters using data from all timepoints (2y–5y), enabling us to characterize longitudinal dynamics and track how individual children transition between clusters over time. We will also examine shorter windows (e.g., 2y-3y, 3y-4y, 4y-5y) and early vs. late periods (2y–3y vs. 4y–5y) to identify short-term or developmental shifts. Clustering can be done by approaches such as Dirichlet multinomial mixtures, or latent class mixed models.^70^ Cluster membership and transitions will be evaluated as candidate predictors of future caries.

Utilising these analyses, we will explore building single-timepoint prognostic prediction—for example, using the oral microbiome at 2y to predict caries experience at 3y. For each outcome type, we will train cross-validated predictive mixed-effects models using the relative abundance of the overall microbiome, or the top-ranking important taxa, as described in D2. This process can be repeated across all timepoints to identify which microbiome features best predict future caries. Finally, to determine which bacterial taxa at earlier timepoints (2y, 3y, 4y) best predict caries experience at a future timepoint, e.g. 5y, we will apply sPLS-DA at each timepoint separately.^43,47^ Resampling, modelling, and optimal model selection will follow the procedures outlined in earlier sections.

###### P3-Combination of questionnaire and oral microbiome data

We aim to improve our prognostic model previously developed in sections P1 and P2, by incorporating I2C longitudinal early-life and oral microbiome data to predict caries in young children at subsequent timepoints. To integrate microbiome data with early-life data, we will employ advanced computational approaches, including smoothing splines within a linear mixed-effects modelling framework.^71^ This approach enables modelling of microbial profiles across sample groups while leveraging sparse multivariate ordination methods. Such ordination methods facilitate the identification of variable sets strongly associated across questionnaire and microbiome datasets, as well as over time.^71,72^

First, we will evaluate early-life factors (e.g., toothbrushing frequency, water fluoridation) to establish an initial predictive framework. Because multiple timepoints are available for both early-life and microbiome data, analyses will be iterated across these timepoints to identify the optimal predictive window. Considering the approaches mentioned in sections P1 and P2, we will then build separate models with and without microbial biomarkers and compare their performance using standardized metrics to assess the added predictive value of the microbiome component.

#### (3) External validation

We will consider external validation by applying the final prediction models to the other cohort (i.e., the model developed from the MIS BAIR will be validated with the I2C, and vice versa). To determine whether external validation is feasible, we will evaluate the availability and comparability of final predictors at specific timepoints—whether data collection schedules and outcome measurements overlap across the two cohorts— and then attempt harmonization of predictors where possible. If certain predictors are unavailable or not directly comparable, the use of clinically justified proxy measures will be explored. Additionally, we will explore the possibility of performing internal-external validation by partitioning our cohorts into clusters based on a specific variable and then iteratively we will use one cluster as the test set while training the model on the remaining clusters.

These feasibility assessments will also guide the decision to pool cohorts to increase sample size and enhance generalisability.

## DISCUSSION

### Strengths

The strengths of this study are manifold. (1) We have access to two rare prospectively collected datasets from early childhood. (2) The availability of longitudinally collected data in both cohorts, obtained through structured questionnaires during the first years of life. This provides rich contextual information on behavioral and environmental factors that may contribute to caries risk, supporting a comprehensive risk assessment framework. (3) Furthermore, the inclusion of our cohort (I2C) where biological samples and dental clinical data are collected longitudinally presents a unique opportunity to compare predictive models built on different types of data. This will help determine whether integrating longitudinal biological sample data enhances predictive accuracy and whether simpler models relying on early-life questionnaire data remain effective. (4) The analysis involves two cohorts representing different populations, with sample sizes starting at 219 participants in the MIS BAIR cohort and an anticipated 400 participants in the I2C cohort. This enhances the robustness and generalisability of the findings. (5) The use of gold-standard dental caries measurement based on ICDAS, rather than self-reported data or other less accurate markers. The use of the ICDAS index provides a validated, standardized, and detailed assessment of caries at various stages, ensuring objective and precise evaluation while minimizing recall bias and misclassification. (6) The substantial overlap in independent variables across cohorts. This uniformity minimizes heterogeneity in outcome assessment and reduces measurement bias. (7) Both studies recruited participants from population health settings rather than from clinical cohorts or specialized health centers, which can minimize selection bias and improve generalisability.

### Limitations

(1) While longitudinal data from critical early life timepoints are available, the MIS BAIR cohort lacks oral microbiome data from the first three years of life. However, the inclusion of the I2C cohort, with fully longitudinally collected biological samples and dental data, helps mitigate this limitation by allowing for comparative analyses. (2) Some differences in early-life data collection through questionnaires across cohorts remain, potentially introducing heterogeneity and affecting model comparability. However, this can be partially resolved through comparable data engineering, i.e., variable building and selection, to harmonize predictor variables across datasets. (3) While external validation is possible, validation in an independent cohort, outside of the research setting, particularly in terms of socioeconomic status (SES), is beyond the scope of this study. However, the insights gained from this analysis will lay the groundwork for developing practical tools that can facilitate extended validation studies across Australia.

## CONCLUSION

Despite progress in caries control technologies, ECC remains a substantial global health challenge. This study aims to develop a new microbiome-based prediction tool that could be used in healthcare settings, to identify high-risk children, before the onset of irreversible clinical disease (cavities). This enables timely, intensive prevention based on accurate and individualized risk prediction.

This prediction tool could potentially (1) enable health professionals to screen for childhood caries and make appropriate referrals, enhancing early intervention and prevention efforts; (2) be incorporated in multi-faceted oral health promotion interventions (application of fluoride varnish or provision of dental care to mothers during pregnancy for identified middle-, high-risk children); and (3) be developed as a chair-side screening kit.

## Ethics statement

This study has ethical and governance approval from The Royal Children’s Hospital Melbourne Human Research Ethics Committee (HREC Reference Number: HREC/111803/RCHM-2024-RCH HREC Reference Number: 111803).

There will be no direct participant involvement in this study. This study will involve the analysis of data from existing data sources. Explicit consent was obtained for the future data analysis of the dental examination.

## Author contributions

YK was responsible for conceptualizing the study, developing the research questions, study design, protocol development and manuscript drafting. MS, SD, KL, and SK contributed to supporting study conceptualization and development of the research questions, protocol, and study design as members of the research team. YK and SK contributed to developing the statistical analysis plan. CB helped draft the microbiome data pre-processing and laboratory section of this study. All authors provided feedback and comments on the drafts and have read and approved the final version.

## Data availability

The datasets generated and analysed during this study are not publicly available due to consent not being obtained from participants for public sharing of data but are available from the corresponding author on reasonable request and conditional on the scope of ethically approved research related to BCG, allergy, infections or child health in the MIS BAIR study.

## Acknowledgements

We thank the Melbourne Infant Study Bacille Calmette Guérin for Allergy and Infection Reduction and INFANT and I2C participants and their families for participating in the dental assessments. We thank Casey Goodall and Nicole Biggs as the data coordinators of the MIS BAIR and I2C cohorts respectively. We thank all the dental examiners of cohorts for their support with data collection. Figures were created with BioRender.

