## Supplementary Material File for "Development and validation of diagnostic and prognostic prediction tools for dental caries in young children: A protocol"

### A- Identification of potential predictor variables

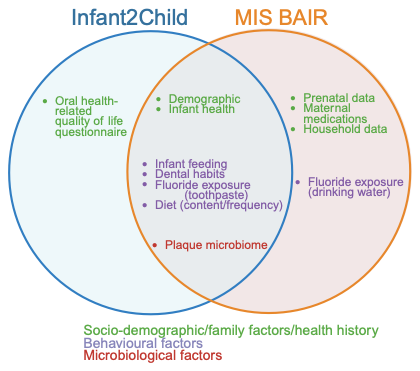
Figure A1. Predictor variables per cohort (Created in BioRender. Khazaei, Y. (2025) <https://BioRender.com/0flh4za>)

#### Sampling procedure and storage media

Study participants are asked to refrain from brushing their teeth the evening before and the morning of sample collection. Sterile swabs (FLOQSwab^TM^, Copan) are used to take the dental plaque samples. For samples collected by parents at home, a plaque home collection kit is used to collect and preserve bacterial DNA prior to batch extraction and processing. Plaque samples collected at home are stored in a viability-preserving microbiostatic medium (VMG II). The medium contains 1% gelatin, 0.05% tryptone, 0.05% peptone P, 0.05% cysteine hydrochloride, 0.05% thioglycolic acid, 1% bacteriological charcoal with a 10% salt solution (0.003% phenyl-mercuric acetate, 0.24% calcium chloride hexahydrate, 0.42% potassium chloride, 1% sodium chloride and 10% sodium glycerophosphate) with 0.5% agar at a pH of 7.3. An aliquot of 1 mL is stored in 2 mL screw-top tubes [1].

For at-home collection, parents are asked to wash their hands, position their child in a comfortable position, hold the swab at the base and run the swab tip along the gumline of the upper and lower teeth. The plaque is immediately placed in 1 mL of the semi-solid VMG II storage medium and posted on the same day. Upon receipt, samples are frozen at -80°C.

Sample collection conducted by a dental practitioner onsite at MCRI involves the same procedure, but samples are placed into an empty collection tube and placed directly into dry ice.

#### Microbiology laboratory procedures and bioinformatics

Raw FASTQ files will be pre-processed for quality check and trimming via pipelines provided in platforms such as Nephele [2]. The DADA2 pipeline will be used for producing Amplicon Sequence Variants (ASVs) and assigning taxonomy to the output sequences [3]. Next, we will convert the data into a phyloseq object and remove non-bacterial sequences (such as mitochondrial reads). Within the DADA2 workflow, the Human Oral Microbiome Database (HOMD) will serve as the reference for assigning taxonomy to the ASVs [4]. Taxonomic assignments will be made based on specific percentage identities: 98% similarity for species-level classification, 95% for genus, and 80% for phylum. To determine the relative abundance of each bacterial taxon, the number of sequencing reads corresponding to a particular species will be divided by the total reads from all species in a sample [5]. The resulting files from both cohorts will then be used for bioinformatic and further statistical analyses.

### B- Outcome

#### Clinical examination

Visual examinations by dental professionals onsite at MCRI are conducted with participants in a semi-supine position on an adjustable examination bed. The examiner stands either beside or behind the participant to optimize access and visibility. A portable LED light (220 lumens; NÄVLINGE clamp spotlight, IKEA) and a dental mirror are used to maximize illumination of the oral cavity, without the use of optical magnification. Plaque collection is conducted immediately prior to the dental assessment. Cotton rolls, gauze, and Puritan swabs are used to dry the teeth, as compressed air is not available.

At-home dental examination can be conducted concurrently with the plaque collection, with the child positioned as per the clinical judgement of the examiner. Dental practitioner will examine teeth present, and the assistant will record dental assessment on the hard copy paper recording sheets including ICDAS. Universal infection control protocols, including additional COVID-19 precautions, are followed throughout all examinations.

The following table describes the criteria used for clinical visual examination and corresponding code. The first digit indicates restorations/sealants—ranging from 0 to 8—while the second digit represents the severity of the carious lesion—ranging from 0 to 6.

Table B1. Criteria for clinical visual examination

| Code | Restorations/sealants codes (first digit) | Caries codes (second digit) |
| --- | --- | --- |
| 0 | Surface not restored/sealed | No visual defect |
| 1 | Sealant, partial | First visual change in enamel (only when air dried OR restricted to pit/fissure) |
| 2 | Sealant, full | Distinct visual change in enamel (widening of pit/fissure when tooth viewed wet) |
| 3 | Tooth-coloured restoration | Localized enamel breakdown (microcavitation) |
| 4 | Amalgam restoration | Underlying dark shadow from dentine |
| 5 | Stainless steel crown | Distinct cavity with visible dentine (< 50% tooth surface) |
| 6 | Porcelain/gold/PFM crown/veneer/onlay/inlay | Extensive cavity with visible dentine (> 50% tooth surface) |
| 7 | Lost/broken restoration | - |
| 8 | Temporary restoration | - |
|  |  | 96 Tooth surface cannot be examined |
|  |  | 97 Tooth missing due to caries |
|  |  | 98 Tooth missing for reasons other than caries (e.g., exfoliated, trauma) |
|  |  | 99 Unerupted |

### C- Statistical analysis

#### C1- Cleaning of predictor variables

We will initially prioritise predictors with established or suspected causal relationships with the outcome (such as sugar consumption, dental habits, fluoride), however, in the absence of evidence of a causal relationship, we will not exclude any potentially relevant predictors a priori [6]. After evaluating the distribution of all predictors, we will consider excluding those with limited variation across the outcome categories, because they will contribute little to our models unless prior evidence indicates a strong association with the outcome [7]. For instance, if caries outcomes (e.g., the number of children with positive caries status) do not vary substantially across categories of household size (e.g., households with 2–3 members, 4–5 members, or 6 or more members), we may remove it from the list of predictors.

Data exploration

Next, we will evaluate the need for data transformations to address potential nonlinear relationships between predictors and outcomes. This evaluation will include visual inspections (e.g., scatter plots, residual plots) in both univariate and multivariate contexts, as well as exploration of various transformation techniques. For continuous variables with highly skewed distributions, we may consider trimming outliers or applying winsorization (e.g., setting extreme values to the 95th percentile). Additionally, we will consider more advanced approaches, such as generalised linear models (GLMs) and generalised additive models (GAMs), which give us more flexibility in modelling data with a skewed distribution, also splines and polynomial models to improve data distribution and model fit [8]. We will always take into account how predictor variables have been modelled in prior prediction models; for instance, categorising breastfeeding practice into exclusive and non-exclusive breastfeeding predictors per the World Health Organization (WHO) [9, 10], and where possible, from previous MIS BAIR cohort publications [11, 12].

#### C2- Creating new predictors

We will use feature engineering to develop new predictor variables, including interaction terms or composite measures, in order to capture more complex relationships between predictors and outcomes over time [8]. For instance, a sugar consumption predictor might be derived from proxy variables such as fruit juice and cake/biscuit intake during the first year of life in the MIS BAIR study and then categorized according to the WHO’s guideline of keeping free sugar intake below 5% of total energy [13]. We will generally avoid dichotomising or categorising continuous predictors, unless necessary, as it reduces information and diminishes statistical power [6]. In the presence of different definitions for one predictor, we will compare the overall predictive performance of the models and the one which improves the overall model fit the most with the highest biological relevance will be chosen for the final model. Additionally, we will consider grouping related predictors—such as various dental habits—into a single predictor at one or multiple timepoints, clustering biomarkers predictive of caries, or combining certain food items into food groups.

#### C3- Predictor variable selection

Discriminant analysis will help identify the key factors (e.g., sugar intake, oral hygiene habits, microbiome markers) that most effectively classify individuals into risk groups, while elastic net regression will refine the selection of predictors and improve the robustness of the model. For discriminant analysis, rigorous verification of underlying assumptions—specifically, multivariate normality of predictor variables and homogeneity of variance-covariance matrices across groups—is critical to ensure model validity and reliable performance outcomes [14, 15]. These complementary methods ensure both accurate classification and optimal predictor variable selection, enabling the development of a reliable prediction tool.

#### C4- (D1) Diagnostic prediction model for caries in young children utilising questionnaire-based data:

In the ten-fold nested CV approach, we will first randomly separate our training (90%) and validation datasets (10%). We will perform parameter tuning and predictor variable selection by using the training dataset and evaluate on the remaining validation test. This process will be repeated 10 times, cycling through all the folds so that each can be the testing set. The overall model’s performance will be calculated as a mean of performance metrics of the 10 separately developed models on different 10% sets of the validation data which will be held out from training. We will consider partially nested CV and bootstrapping as other alternatives.

Traditional statistical methods such as logistic regression, as a parametric model, rely on specific assumptions (e.g., independence of errors, linearity in the logit for continuous predictor variables, absence of multicollinearity) and emphasize hypothesis-driven analyses [16]. These methods generally favour simpler, more interpretable models over complex ones. In contrast, machine learning algorithms are effective in analysing complex, high-dimensional data, particularly in the presence of complicated nonlinear interactions among predictors. As non-parametric approaches, they impose minimal assumptions about the underlying data-generating process, offering greater flexibility in modelling complex relationships [16]. Random Forest builds an ensemble of decision trees, leveraging bootstrapped sampling and random feature selection to enhance model robustness and reduce overfitting [17]. In contrast, gradient boosting sequentially builds an ensemble by iteratively reducing prediction errors, yielding a model that can capture intricate data structures and subtle interactions among predictors [18]. These machine learning approaches offer high predictive power, which may improve the accuracy and reliability of the caries prediction tool. However, despite their convincing predictive performance, the lack of an explicit model structure makes it challenging to directly correlate machine learning-driven results with existing biological knowledge [19].

#### C5- (D2) Diagnostic prediction model for caries in young children utilising oral microbiome data:

Diversity analyses include alpha and beta diversity analyses where beta diversity quantifies the dissimilarity between communities (multiple samples), as opposed to alpha diversity, which focuses on variation within a community (one sample). We will then analyse the statistical significance of alpha and beta diversities using different methods [3, 20, 21]. We will begin by examining the number of reads (i.e., library size) in each sample to detect any substantial variation, using the R packages ggplot2 and gghighlight for visualization. Next, we will calculate species alpha diversity with the phyloseq package, employing either rarefaction or normalization to a median sequencing depth (depending on the rarefaction curves) [22]. Our alpha diversity metrics include observed species richness, the Shannon index (encompassing both richness and evenness), and the inverse Simpson index (also reflecting richness and evenness but typically less sensitive to rare taxa compared with the Shannon index) based on all species. We will examine if using 99% similarity as the cutoff for species identification, as opposed to 100% or 97%, is necessary to account for potential nucleotide changes introduced during amplification. We will visualize alpha diversity using boxplots generated in ggplot2 and ggpubr. For comparisons of relative abundance results, α-diversity results and statistical significance between caries groups, we will use the non-parametric Wilcoxon or Mann-Whitney test, due to their flexibility. Subsequently, we compute beta diversity measures at the species level—such as Bray–Curtis dissimilarity and Unweighted UniFrac distances—using non-rarefied data, and restricted to species with a minimum relative abundance of 0.001 and minimum prevalence of 10% [5]. Other compositional beta diversity such as Information UniFrac will also be considered [22, 23].

There are many methods to conduct Differential Abundance Analysis (DAA) and we will consider performing them to ensure robust outcomes [23]. By this, we will gain important insights into the underlying mechanisms of caries.

### D- Patient and public involvement

MIS BAIR did not have formalized processes in place for consumer or public involvement in the design, conduct, reporting, or dissemination of the research. However, participants were provided with an opportunity to offer feedback at the end of each survey regarding their experience in the study. This input informed modifications to the study design. The Infant2Child cohort study includes engagement with consumer representatives, who meet regularly to inform project process.
